# Development, Testing, and Calibration of LINCS: A New Microsimulation Model of Maternal and Fetal Cytomegalovirus Infection in the US

**DOI:** 10.1101/2024.09.30.24314653

**Authors:** Aaron S. Wu, Elif Coskun, Malavika Prabhu, Emily M. Santos, Fatima Kakkar, Clare F. Flanagan, Caitlin M. Dugdale, Megan Pesch, John C. Giardina, Andrea L. Ciaranello

## Abstract

**Background:** Congenital cytomegalovirus (cCMV) is a leading cause of birth defects and the most common cause of non-genetic sensorineural hearing loss in children. There is a lack of decision modeling frameworks that can project cytomegalovirus (CMV)-related patient outcomes and inform health policy. We created, tested, and calibrated a model of CMV acquisition and transmission in pregnancy using linked mother-infant dyads.

**Methods:** We developed the **L**inking **IN**fants and Mothers in **C**ytomegalovirus **S**imulation (LINCS) dyad-level Monte-Carlo microsimulation model of CMV infection among pregnant people and fetuses throughout pregnancy. We parameterized the model with data from the US, Canada and the EU from the existing literature, implemented rigorous code testing procedures, and calibrated a key set of parameters to match model output to external data on cCMV prevalence and symptom risk.

**Results:** A fully parameterized model for CMV among pregnant people and fetuses was developed, and the model code was confirmed to perform as specified. The calibration procedure identified parameter sets that generated model output closely matching the target values from the available data on cCMV prevalence and symptom risk.

**Conclusions:** The LINCS model’s ability to simulate the natural history of CMV infection during pregnancy was described and demonstrated, and the model was tested and calibrated to ensure proper functioning. Base case parameters were derived for CMV infection natural history to be used in future decision analyses of CMV testing and treatment strategies.

## INTRODUCTION

Congenital cytomegalovirus (cCMV) is the most frequent infectious cause of birth defects and the leading cause of non-genetic sensorineural hearing loss in newborns and infants.^1^ Pregnant people are often exposed to and infected with CMV during pregnancy (“maternal infection”), frequently through contact with saliva or urine from children in daycare or preschool.^2^ CMV infections in adults often present with minimal or no symptoms, leading to frequently missed diagnosis and treatment of infection during pregnancy, and increasing the risk of birth defects or other fetal complications. It is estimated that 0.3-0.6% of all newborns in the US are born with cCMV, which in 2022 would have been between 10,000 to 22,000 cCMV cases in newborns.^3^ Of infants with cCMV, up to 30% are born with or will go on to develop cognitive impairment, vision loss, cerebral palsy, or hearing loss.^3,4^ In addition to these health impacts, the lifetime additional costs of care for an infant with symptomatic CMV is estimated to exceed $1.3 million.^5^

Despite the major health and economic burdens of cCMV, the best practices for preventing, diagnosing, and treating CMV in pregnant people and infants remain uncertain.^6^ Interventions to reduce risks of maternal infection, vertical transmission, and disease severity among affected newborns have recently emerged,^7^ but the lack of accurate diagnostic tools and the uncertain benefits of treatment pose challenges in the prevention and management of cCMV. Some organizations like the European Society for Clinical Virology-European Congenital CMV Initiative (ESCV-ECCI) and the Society of Obstetricians and Gynaecologists of Canada (SCOG) recommend screening for CMV serostatus among all pregnant women.^8,9^ However, current guidelines from major US professional societies do not recommend universal cCMV screening or treatment in pregnancy, primarily due to concerns about imperfect diagnostic tests and limited confidence in trial data supporting novel therapies for the treatment of maternal or fetal CMV infection.^10,11^

Disease simulation modeling can synthesize the best available data from multiple sources, project long-term outcomes beyond the horizon of clinical trials or cohort studies, evaluate multiple strategies simultaneously, and explicitly evaluate the tradeoffs in benefits and harms.^12–15^ Modeling can add substantial value to traditional study designs such as trials or observational cohorts and can inform the development of clinical care guidelines.^16–18^ However, there are limited published models of CMV infection that use discrete-timestep agent-based microsimulation methods, with previous studies mostly focused on vaccination.^19–21^ Previous models capable of simulating CMV and cCMV screening strategies primarily use deterministic or probabilistic state-transition decision trees.^22–26^ To our knowledge, no agent-based discrete-time microsimulation models have been developed to simulate such a wide array of testing strategies or emerging therapies for maternal, fetal, or infant cCMV. Agent-based microsimulation (ABM) modeling has the advantage of allowing for more agent-specific heterogeneity than with cohort-based models, and the discrete-time structure allows for explicit control over the timing of events.^27^ There are very few models focusing on CMV acquisition during pregnancy in the United States setting.^22,23^ These Markov model-based studies showed that prenatal screening during the first trimester for CMV would be cost-effective if treatment with valacyclovir followed a diagnosis of a primary maternal infection in the first trimester.^22,23^ Unlike these previous models of CMV testing and treatment during pregnancy in the US, the LINCS model also accounts for non-primary infection (reinfection or reactivation), which can, in theory, affect the utility and therefore cost-effectiveness of maternal CMV tests, as risks of vertical transmission may be different for non-primary infections. Although non-primary infection is frequently described as conferring markedly lower risk of vertical transmission, empirical data on differences in transmission dynamics between primary and non-primary infection are very limited.^28,29^ The model allows us to perform calibration and sensitivity analyses on this uncertain parameter. The discrete timestep nature also allows for the simulation of serologic testing-based algorithms, such as IgG avidity, which can approximately estimate timing of infection. Since key cCMV outcome risks can be impacted by type (primary vs. non-primary infection) and timing of infection, identifying infection type and timing is important to clinical decisions. This model structure allows us to simulate how different testing algorithms interact with the kinetics of viral load and serologic response throughout the course of pregnancy.

Our objective was to calibrate CMV epidemiological parameters to US data and biological behavior of the model as a foundation to conduct future prenatal CMV screening and treatment analyses and project clinical outcomes of CMV infection during pregnancy, primarily within the US but amongst other populations of interest as well.

## METHODS

### Overview

We developed the Linking INfants and Mothers in Cytomegalovirus Simulation (LINCS) microsimulation model to project clinical outcomes and costs among pregnant people at risk for CMV infection during pregnancy and their infants. The LINCS model can be used to evaluate current and novel testing and treatment strategies in decision analyses, including cost-effectiveness and comparative effectiveness analyses. Cohort characteristics in the model can be adjusted to reflect the epidemiological dynamics and demographic characteristics of different populations. We derived model input parameters from cohort studies and clinical trials, including rates of non-primary CMV infection in pregnancy, accuracy of diagnostic tests for CMV, and effectiveness of currently available treatments (Table 1). We also used a Bayesian calibration approach to estimate key uncertain model inputs – we used published estimates of neonatal cCMV prevalence and symptomatic cCMV risk in North American settings as calibration targets and calibrated 6 model parameters, including rates of primary (first-ever) CMV infection in pregnancy and risk of vertical transmission following primary and non-primary infection. We verified the code used to implement the model structure and checked for errors by comparing model outputs to the data used as model inputs, evaluating the model at extreme parameter values, and inspecting traces of individual patient clinical trajectories. Additional code quality testing strategies were performed to verify that each part of the model and the interactions between them were behaving as specified.

**Table 1.** Selected data inputs for the LINCS model of CMV in pregnancy.

| Input | Value | Source |
| --- | --- | --- |
| **Cohort** |  |  |
| Maternal age, mean (SD) | 26 (5) | 56 |
| Maternal primary CMV incidence rate, per week, % | Included as calibration parameter; final calibrated values shown in Table 3 |  |
| Maternal CMV seroprevalence, % | Calculated based on calibrated primary CMV incidence rate; final calibrated values shown in Table 3 |  |
| Probability of vertical transmission (by maternal infection type and timing), % | Included as calibration parameter; final calibrated values shown in Table 3 |  |
| Proportion of cCMV that is symptomatic at birth, by timing of maternal infection, % | Included as calibration parameter; final calibrated values shown in Table 3 |  |
| Weekly probability of premature birth, range by maternal age, % | Weeks 24-31:<br>Age <24: 0.7%<br>Age 25-29: 0.6%<br>Age 30-34: 0.6%<br>Age 35-39: 0.8%<br>Age >40: 1.2%<br>Weeks 32-36:<br>Age <24: 4.7%<br>Age 25-29: 4.1%<br>Age 30-34: 3.9%<br>Age 35-39: 4.4%<br>Age >40: 5.2% | 65 |
| Probability of fetal demise, range by maternal age in years, % | Before 20 weeks (miscarriage):<br>Age <20: 16.2<br>Age 20-24: 11.2%<br>Age 25-29: 9.7%<br>Age 30-34: 10.8%<br>Age 35-39: 16.7%<br>Age 40-44: 33.2%<br>Age >45: 56.9<br>At/after 20 weeks (stillbirth):<br>Age <20: 0.2%<br>Age 20-24: 0.2%<br>Age 25-29: 0.3%<br>Age 30-34: 0.3%<br>Age 35-39: 0.4%<br>Age 40-44: 0.3%<br>Age >45: 0.3% | 66 |
| **Baseline prenatal visit schedule** |  |  |
| Obstetric visit frequency, by trimester | Weeks 6-28: 1/month<br>Weeks 29-36: 1-2 weeks<br>Weeks 36-39: 1/week | 67 |
| Timing of first anatomic scan ultrasound | 20 weeks | 67 |
| Testing |  |  |
| Assays to detect maternal primary CMV infection during pregnancy |  |  |
| Serum IgM sensitivity / specificity, % | 90 / 96 | 68 |
| Serum IgG sensitivity / specificity, % | 97 / 96 | 69 |
| Serum IgG avidity sensitivity / specificity % | 94.3 / 100 | 70 |
| Serum CMV DNA PCR sensitivity / specificity, % | 94 / 99 | 71 |
| Assays to detect fetal cCMV infection |  |  |
| Amniotic fluid CMV DNA PCR sensitivity / specificity, % | 86 / 100 | 39 |
| Probability of follow-up testing after abnormal ultrasound result, % | 60-100 | Assumption |
| Probability of follow-up testing after abnormal IgM-IgG result, % | 95-100 | Assumption |
| Therapies |  |  |
| Valacyclovir for primary infection in periconception and first trimester, odds ratio of vertical transmission risk | 0.34 | 43 |
| CMV: cytomegalovirus; Ig: immunoglobulin; PCR: polymerase chain reaction; SD: standard deviation. |  |  |

### Model structure

#### Mother-infant dyads

The LINCS model is a dyad-level Monte-Carlo microsimulation model with a weekly time step that begins for each mother-child pair at conception and ends at delivery. All patients are modeled as a linked mother-child dyad throughout pregnancy and delivery and enter the simulation at the time of conception (week 2 of gestational age). The representation of the mother and child as linked allows for more detailed simulation of both biological and clinical interactions between the mother and fetus during pregnancy, and helps account for the interactions between different prevention, screening, and treatment strategies in decision analyses. A literature review found that previous microsimulation models of CMV in pregnancy modeled the mother and child separately, which limited the ability of those models to assess interactions between strategies involving both the mother and child.^24,26,30,31^

#### Maternal CMV infection

At model start, all pregnant people are assigned an age and previous CMV infection history. CMV infection history includes infection that occurred <u>remotely before conception</u> (for this analysis, >3 months before conception), infection that occurred <u>recently preconception</u> (for this analysis, ≤3 months to 3 weeks prior), infection that occurred <u>periconception</u> (within 3 weeks prior to conception), or <u>no prior CMV infection</u>. This differentiation enables the model to account for clinical and diagnostic implications of type and timing of CMV infection (and can be varied as needed). For example, this structure can be used when assessing serologic tests that do not reliably distinguish between infection 3-12 months prior to conception (conferring no risk to the fetus) or <3 months (when risk of cCMV is present, conferring a substantial risk to the fetus).^28,32,33^

Pregnant people with a recent preconception or periconception infection are assumed not to be at risk for another maternal CMV infection during pregnancy. Although data on strain-specific immunity are sparse, we assume a 12-month duration of protection from reinfection.^34^ All others face a weekly probability of CMV infection. Those with remote preconception infection are at risk for non-primary CMV infection (reinfection or reactivation); those with no prior CMV infection are at risk for primary infection (Table 1).^33^

#### Vertical transmission

The risk of vertical transmission (VT) to modeled fetuses depends on the type and timing of maternal CMV infection. With recent preconception or periconception maternal infection, simulated fetuses face a risk of VT modeled to occur at week 6, based on expert opinion that sufficient embryonic development has occurred to permit transmission.^35–37^ For maternal infection that occurs during pregnancy (either primary or non-primary infection), modeled fetuses face a one-time risk of VT 6 weeks after maternal infection occurred, allowing time for maternal viremia, placental infection, and fetal infection. VT risks are lower for maternal infections that occur in the first trimester compared to later in pregnancy, although there is a higher risk of symptomatic disease if cCMV does occur following first trimester maternal infection. VT risks are higher for primary compared to secondary maternal infections.

The model divides congenital CMV (cCMV) into five mutually exclusive “phenotypes” to reflect multiple possible manifestations of cCMV for infants/children, including 1) fully asymptomatic cCMV infection at birth and for life, 2) transient laboratory abnormalities or fetal growth restriction (FGR) at birth, with no lifelong symptoms, 3) hearing loss present at birth or developing later; these infants may also develop vestibular dysfunction (impaired balance and gait), speech and language delay, or sensory integration disorders, 4) mild to moderate neurodevelopmental delay; these infants may have mild clinical symptoms (hypotonia, hepatosplenomegaly) and/or nonspecific neuroimaging findings, and may develop sensorineural hearing loss (SNHL), and 5) severe neurodevelopmental delay (e.g., severe autism); these infants are non-ambulatory and/or non-verbal lifelong, and may have extensive abnormalities on neuroimaging, clinical signs of severe cCMV (e.g., microcephaly, severe FGR), with or without SNHL and/or severe laboratory abnormalities (e.g., thrombocytopenia, neutropenia, jaundice).^2^ For this analysis, we populated phenotypes based on visible symptoms at birth, in two categories: asymptomatic (1 and 2) and symptomatic (3, 4, and 5), although the model program is flexible to simulate additional phenotypes.^33^

#### Diagnostic testing

Maternal CMV and fetal cCMV can be diagnosed via user-specified prenatal assays. Tests to detect <u>maternal CMV infection</u> include serum assays such as CMV DNA polymerase chain reaction (PCR), IgM antibody, IgG antibody, and IgG avidity (feasible to assess when IgG is present; greater avidity of the IgG antibody to CMV antigens reflects longer time since infection occurred).^28^ We derived the sensitivity and specificity for each of these diagnostic tests from the published literature (Table 1). In the model, these sensitivity and specificities are applied alongside modeled viral load and serology kinetics in pregnant people (Figure 2). For example, following maternal infection, CMV viremia is modeled to occur after 1 week and resolve by 3 weeks; CMV DNA PCR can detect this with the listed sensitivity. Detection of CMV DNA in maternal serum is generally considered diagnostic of maternal CMV infection. IgM antibody is modeled to develop within 2 weeks and persist for a mean of 52 weeks after infection (standard deviation [SD] 4 weeks). IgG is modeled to develop within 3 weeks and persist lifelong. IgG avidity is modeled as “low” when IgG first develops and rises to “high” at 22 weeks (SD: 2 weeks) after infection, reflecting the cutoff values and estimated timing from commercial assays.^38^

If VT has occurred, tests that may identify cCMV infection in the fetus include routine diagnostic ultrasound (R-US), detailed diagnostic ultrasound (D-US), and amniocentesis (AC) followed by CMV DNA PCR performed on amniotic fluid.^28^ The sensitivity and specificity of these tests are also applied alongside biologic events in the fetus.^39^ Following VT, infant infection can lead to impaired embryogenesis or fetal development, resulting in abnormalities detectable on ultrasound after 18 weeks of gestational age.^40^ These abnormalities are not specific to CMV and can occur with many other conditions, and they also do not occur in all fetal cCMV infections, creating imperfect sensitivity and specificity of ultrasounds for CMV.^41^ CMV DNA may become present in amniotic fluid (which is comprised largely of fetal urine) after 20 weeks of gestation, allowing for sufficient fetal renal development to permit excretion of DNA in fetal urine, and persists until delivery.^39^ Detection of CMV DNA in amniotic fluid is generally considered to confirm fetal infection.^42^

The model user can specify the proportion of primary and non-primary maternal infections that are symptomatic, and the proportion of symptomatic maternal infections that are diagnosed as CMV. The user can also specify diagnostic testing algorithms to reflect current clinical care practices or novel diagnostic algorithms. For example, IgM/IgG serologic testing can be offered as part of routine screening of all pregnant people, as recommended by ECCI and SCOG, or as part of focused testing following known exposure, maternal symptoms consistent with CMV, or fetal ultrasound findings concerning for cCMV, as is more commonly done in the US (Table 1).^6,9,10^ Amniocentesis with amniotic fluid CMV DNA PCR can be offered following confirmed maternal primary infection or abnormal findings on fetal ultrasonography. These diagnostic algorithms and the clinical interpretation of test results are shown in Table 2.

**Table 2.** Diagnostic algorithms and interpretations for CMV in pregnancy and congenital CMV. ^9^

| IgM | IgG | Avidity | Interpretation | Follow-up test | Follow-up result | Follow-up interpretation |
| --- | --- | --- | --- | --- | --- | --- |
| Negative | Negative | -- | No prior CMV infection | n/a | n/a | n/a |
| Negative | Positive | -- | Remote PI | No follow-up | n/a | n/a |
| Positive | Negative | -- | Recent PI **or** FP IgM | Serum CMV DNA PCR | Positive | Recent PI |
|  |  |  |  |  | Negative | No prior infection, FP IgM |
| Positive | Positive | Low | Recent PI | -- | -- | -- |
|  |  | High | Remote PI, with recent NPI **or** FP IgM | Amniocentesis with amniotic fluid CMV DNA PCR | Positive | Vertical transmission |
|  |  |  |  |  | Negative | No vertical transmission |
| PI: primary infection. FP: false positive. NPI: non-primary infection. |  |  |  |  |  |  |

#### Therapies

We calibrated the model assuming no antiviral therapy during pregnancy, reflecting the settings in which the data used as calibration targets were collected. However, for future policy analyses, the structure of the model permits the user to specify therapeutic interventions in pregnancy following diagnosed maternal or fetal CMV infection. For example, treatment with high dose valacyclovir (8 grams/day) is recommended in some settings following confirmed maternal primary infection in the first trimester with the goal of <u>preventing fetal infection</u>; this is continued through the time of amniocentesis.^8,9^ We will model the effectiveness of this intervention as a reduction in VT risk (Table 1).^43^ Following confirmed fetal infection, high dose valacyclovir can also be offered to a pregnant woman to <u>treat fetal infection</u> and reduce the severity of infant cCMV disease.^9,43,44^ Although there is no randomized controlled trial evidence currently available to parameterize the effectiveness of this treatment on cCMV severity, the model structure permits sensitivity and scenario analyses on fetal treatment by adjusting the phenotype distribution for newborns with cCMV, shifting the distribution under treatment toward milder, less symptomatic cCMV phenotypes.

#### Model outcomes

In each week of the simulation, the model tracks true maternal CMV infection status, true fetal cCMV status, true biomarker status (e.g., IgM, IgG, DNA presence in maternal serum; DNA presence in amniotic fluid), and fetal survival. At the end of each week, there are three possible pregnancy-related health states: continuation of the pregnancy into the following week, live birth (considered premature if before week 37), or fetal demise (considered spontaneous abortion/miscarriage if before 20 weeks, and intrauterine fetal demise/stillbirth if after 20 weeks; probability depends on true fetal cCMV status and week of gestation). If neither live birth nor fetal demise occurs, the pregnancy continues into the next week (Figure 1). After birth, a user-specified proportion of infants with hearing loss or visible symptoms will be diagnosed with cCMV, and a probability of immediate neonatal death depends on true cCMV status and the week of gestation at which birth occurred. After the entire cohort is simulated, summary statistics are tallied, including number of live births, number of preterm deliveries, neonates with cCMV, distribution of neonatal cCMV phenotypes, number of maternal infections diagnosed during pregnancy, and number of neonates diagnosed with cCMV.

**Figure 1.**
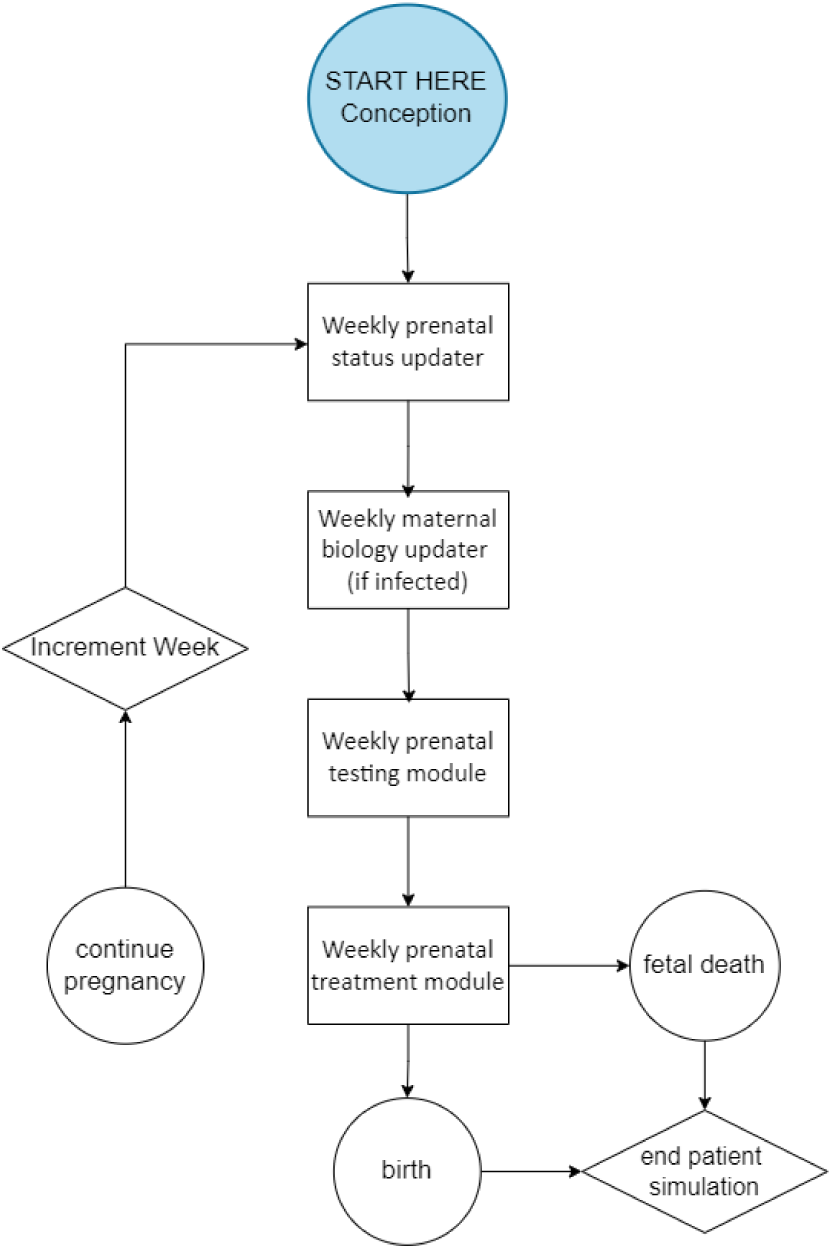
Schematic diagram of the structure of the LINCS model.

**Figure 2.**
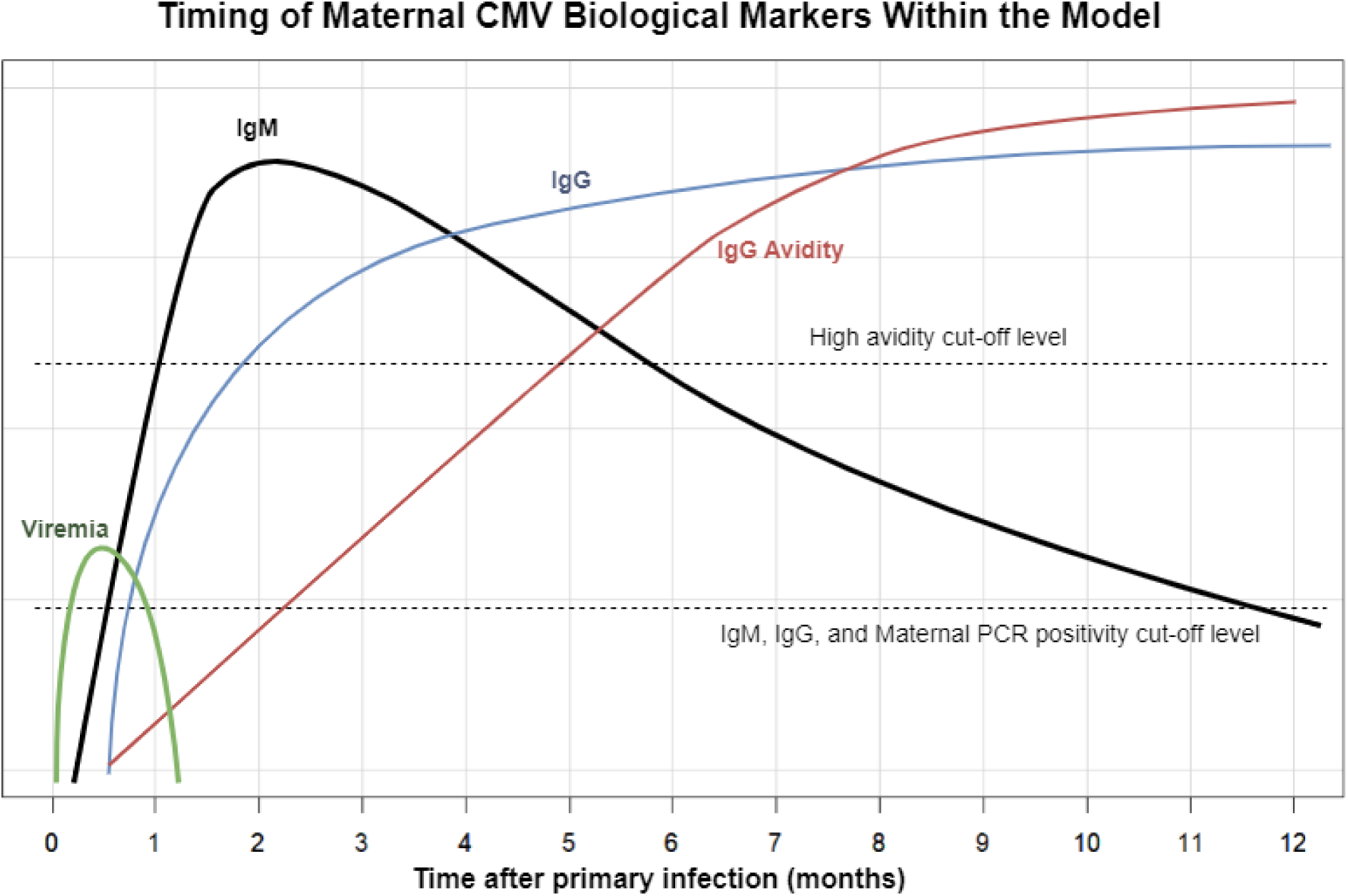
Timing of Maternal Biological Markers in Primary CMV Infection in the LINCS model. Base-case representation of changes in anti-cytomegalovirus (CMV) IgM (immunoglobulin M) and IgG (immunoglobulin G) antibody levels, as well as IgG avidity, following a primary CMV infection.^28,38^ IgM production occurs during both primary infection and reinfection, and can persist in some cases for more than a year following an acute primary infection.^28^ IgG antibodies persist for life following acute primary infection, and IgG avidity gradually increases during the months following primary infection. Maternal CMV viremia is detectable by PCR testing between 1 and 3 weeks after infection in the base case.^34,64^ This timing of when each biological marker meets the threshold for positivity can be defined by the model user.

### Unit testing and functional testing of model dynamics

Unit testing and functional testing are code and software development methodologies to ensure code quality and correct function in accordance with specifications. Testing was performed at each step in the development of the simulation model to ensure code quality and proper function. Descriptions of these methods can be found in the supplemental materials.

### Model calibration

#### Overview of calibration approach

We implemented a Bayesian calibration approach to calibrate five groups of key parameters in the model.^45^ Our two calibration targets were the observed cCMV prevalence in a universal screening program in Minnesota and the observed fraction of cCMV cases that are symptomatic in universal screening programs in Minnesota and Ontario.^46,47^ The five calibrated parameters were: (1) *rate of maternal infection and maternal seroprevalence*, (2) risk of VT following maternal primary infection (*VT:primary*), (3) relative risk of VT following non-primary maternal infection (*RR-VT:non-primary*) compared to risk following primary infection, (4) proportion of cCMV that is symptomatic at birth following maternal primary infection (*symptomatic:primary*), and (5) relative proportion of cCMV that is symptomatic at birth following maternal non-primary infection (*RR-symptomatic:non-primary*) compared to symptomatic cCMV following maternal primary infection. VT and symptomatic cCMV risks are stratified by timing of maternal infection – preconception (1-3 months prior to conception), periconception (0-1 months prior to conception), 1^st^ trimester, 2^nd^ trimester, and 3^rd^ trimester. Following the guidance outlined by Menzies *et al*. for Bayesian calibration using the sampling importance resampling algorithm, we sampled from prior distributions for each of the parameters, calculated the likelihood of the model output given the observed data for the calibration targets, and then resampled from the original parameter draws with replacement using the calibration target likelihood as sample weights to obtain posterior distributions of the model parameters listed above.^45,48^ We provide a summary of this process below, and a detailed description of the derivation of the prior distributions for the input parameters is included in the supplemental materials.

#### Prior distributions for input parameters

##### Maternal CMV infection incidence rates and seroprevalence

The incidence rates of primary maternal CMV infection were drawn from serology data collected by the National Health and Nutrition Examination Survey (NHANES) from 1999-2004.^49^ To derive age-specific primary incidence rates from the cross-sectional CMV seroprevalence data provided by NHANES (i.e., positive or negative for CMV antibodies on the survey date), we estimated an interval censored hazard regression model, since we know incident CMV infection occurred for individuals with CMV antibodies between birth and their age when the data was collected.^50^ We fit the hazard regression using natural cubic splines for age with 3 knots, implemented using the *flexsurv* package in R.^51^ Based on the estimated hazard model, we also calculated seroprevalence given maternal age at model start. To generate a distribution for the incidence rate that we could use as a prior in the calibration procedure, we used a bootstrap approach that accounted for the complex survey design in NHANES: we generated 10,000 replicate datasets using the *svrep* R package, and re-estimated the hazard model in each replicate to generate a distribution for the maternal incidence rate and seroprevalence estimates.^52^

To estimate the incidence rate of non-primary infection, we calculated the relative risk of non-primary compared to primary maternal infection using results from Paris *et al*.^53^ This prospective study recorded 22 primary and 12 non-primary (defined as >4-fold IgG increase) infections over 36 months out of 149 and 204 adolescent girls who were seronegative and seropositive, respectively, at baseline. A Poisson regression with an offset term was used to estimate a prior distribution for the relative risk of non-primary to primary infection.

##### Vertical transmission risk and symptomatic cCMV infection risk

The priors for *VT:primary* and *symptomatic:primary* were drawn from the results of studies reported by Chatzakis *et al.*^33^ We fit Bayesian binomial regressions with study-level random effects to estimate the *VT:primary* and *symptomatic:primary* risks stratified by the timing of maternal infection using the *rstanarm* R package;^54^ we used the results of these regressions as the priors for the *VT:primary* and *symptomatic:primary* risks. We set the priors for *RR-VT:non-primary* and *RR-symptomatic:non-primary* as uniform distributions between 0-1, based on previous suggestions that VT risk from non-primary infection, and risk of infant symptoms if infection does occur, are lower than from primary infection.^28,29^

#### Calibration targets

##### Neonatal cCMV prevalence

To derive the calibration target for neonatal cCMV prevalence, we used the results from a universal newborn cCMV screening program in Minnesota reported by Kaye *et al.*; out of 60,115 tests, they reported 174 positive results within the first 21 days of life, giving an observed prevalence of 0.29%.^46^ To compare LINCS output to the Kaye *et al.* data, we first adjusted this observed prevalence for the imperfect sensitivity of the dried blood spot (DBS) test used in the Minnesota screening program in the following way: Dollard *et al.* separately reported that the relevant type of DBS-based assay correctly identified 41 out of 56 cases (73% sensitivity), with perfect specificity.^55^ We constructed a likelihood function for the underlying cCMV prevalence in the Minnesota cohort, assuming a binomial distribution with prevalence *p* for the Kaye *et al.* results and a beta distribution for the test sensitivity *s* from Dollard *et al.* (with beta distribution parameters α=42 and β=16): 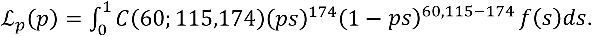 Here, *C(n;k)* is the binomial coefficient, and *f* is the probability density function for the beta distribution. Based on this approach, the maximum likelihood estimate for the underlying cCMV prevalence is 0.394%.

##### Proportion of cCMV that is symptomatic at birth

To derive the calibration targets for the proportions of cCMV infections that are symptomatic at birth, we used results from the Minnesota and Ontario universal screening programs.^46,47^ Kaye *et al.* reported that out of 176 infants with confirmed cCMV infection and/or disease from the Minnesota universal screening program, 21 (12%) had cCMV disease according to the CDC definition.^46^ In a similar newborn screening program in Ontario, Canada, Dunn *et al*. reported that out of 601 infants with cCMV who underwent medical and audiologic assessments, 96 (16%) were symptomatic, defined as “1 or more abnormalities compatible with cCMV and/or SNHL at birth).^47^ Combining these results (117 symptomatic cases out of 777 infections) led to an overall symptom risk of 15.1%. Assuming perfect sensitivity and specificity of symptomatic screening, these results provide the following likelihood function for the model-predicted probability *r* of symptoms at birth among infants with cCMV infection: ℒ_*r*_(*r*) = *C*(777; 117)*r*^117^(1 −*r*)^777−117^.

##### Overall likelihood function of calibration targets

We define the overall likelihood function for the calibration targets as the product of the individual likelihood functions for the prevalence of cCMV and the proportion of cCMV that is symptomatic at birth listed above (i.e., ℒ_*p*_ and ℒ_*r*_(*r*)), assuming independence between each target. This is the function used to evaluate the fit of the model output for the proportion of newborns with cCMV and the proportion of cCMV newborns with symptomatic CMV, when compared to the calibration targets.

#### Calibration runs

We drew 10,000 parameter sets from the prior distributions listed above. We then ran the model for each of these parameter sets; a cohort size of 20 million mother-infant dyads was used (an initial analysis showed that model outcomes were stable across parameters at a cohort size of greater than 15 million). The ages of pregnant people at model start (15 – 49) were drawn from a truncated normal distribution with a mean of 26 years and a standard deviation of 5 years,^56^ and the base case parameter values from Table 1 were used (aside from the parameters being calibrated) with no treatments applied. We simulated only singleton, viable pregnancies for this analysis. All modeled pregnant people were assumed to be otherwise healthy and remain alive throughout the duration of the simulation.

After running the model, we extracted the overall cCMV prevalence at birth and proportion of newborns with symptomatic cCMV for each run and calculated the corresponding likelihood for each of the 10,000 parameter sets using the overall likelihood function discussed above. We then resampled from these parameter sets (with replacement), using the likelihood values as sampling weights to obtain a posterior distribution of the calibrated input parameters. All calculations were conducted in R 4.4.1.

## RESULTS

### Unit and functional testing

Unit and functional testing were satisfactory to ensure the logic and performance of the model; selected input parameters were all accurately reproduced from the model output within 1% of the true value. Extreme value testing and trace file examination verified that the model was performing as expected. These results are reported in Table S1.

### Calibration

The best-fitting parameter sets generated by the calibration procedures are reported in Table 3. Likelihood values from the simulations to calibrate prevalence of cCMV at birth are reported in Table S2. Using the best-fitting parameter sets, the modeled prevalence of cCMV at birth had a mean of 0.403% (compared with the mean target value of 0.394% described in methods above). The mean modeled proportion of symptomatic cCMV was 14.53% (compared with the target value of 15.1% described above).

**Table 3.** LINCS model calibration results.

| **Input Parameters** |  |  |
| --- | --- | --- |
| **Parameter** | **Prior distribution (95% CI)** | **Posterior distribution (95% CI)** |
| Maternal primary infection rate at age 26<br>(*Rate of maternal primary infection*) | 1.95 (1.59, 2.43) cases/100 person-years | 1.93 (1.67, 2.64) cases/100 person-years |
| Maternal seroprevalence at age 26 | 55.8% (52.5, 59.9%) | 55.6% (52.6, 59.8%) |
| Risk of vertical transmission following maternal primary infection, stratified by timing of maternal primary infection (% risk)<sup>a</sup><br>(*VT:primary*) | Recent preconception: 5.5% (2.9-8.8)<br>Periconception: 17.2% (12.3, 22.9)<br>1<sup>st</sup> trimester: 36.9% (31.5, 43.4)<br>2<sup>nd</sup> trimester: 41.3% (35.3, 48.1)<br>3<sup>rd</sup> trimester: 66.2% (58.6, 73.0) | Recent preconception: 5.4% (3.4, 9.0)<br>Periconception: 18.0% (13.3, 25.0)<br>1<sup>st</sup> trimester: 37.6% (30.3, 45.8)<br>2<sup>nd</sup> trimester: 41.6% (36.4, 48.4)<br>3<sup>rd</sup> trimester: 66.5% (59.4, 75.0) |
| Relative risk of VT following non-primary maternal infection compared to risk following primary infection (*RR-VT:non-primary*) | 0.5 (0.025, 0.975) | 0.495 (0.014, 0.968) |
| Proportion of cCMV that is symptomatic at birth following maternal primary infection, stratified by timing of maternal primary infection (% risk)<sup>a</sup><br>(*symptomatic:primary*) | Pre/periconception<sup>b</sup> and 1<sup>st</sup> trimester: 21.8% (12.7, 39.5)<br>2<sup>nd</sup> trimester: 3.4% (1.0, 8.3)<br>3<sup>rd</sup> trimester: 0.6% (0.002, 3.4) | Pre/periconception<sup>b</sup> and 1<sup>st</sup> trimester: 47.3% (34.8, 57.9)<br>2<sup>nd</sup> trimester: 9.9% (4.6, 18.6)<br>3<sup>rd</sup> trimester: 2.0% (0.02, 10.3) |
| Relative proportion of cCMV that is symptomatic at birth following maternal non-primary infection (*RR-symptomatic:non-primary*) | 0.5 (0.025, 0.975) | 0.584 (0.102, 0.961) |
| **Calibration Targets** |  |  |
| **Outcome** | **Empirical Target** | **Model-Estimated Result (95% CI)** |
| Neonatal cCMV Prevalence | 0.394% | 0.403% (0.337, 0.480) |
| Proportion of all cCMV infections that are symptomatic at birth | 15.1% | 14.53% (12.3, 17.2) |
| a. Estimates for prior drawn from binomial regression of results reported by studies listed in Chatzakis et al.<sup>33</sup> Note there are slight differences in priors listed here and meta-analysis results reported by Chatzakis et al.<sup>33</sup> because of differences in analytic approach.<br>b. The pre/periconception and 1<sup>st</sup> trimester time periods are grouped here because studies were only available from Chatzakis et al.<sup>33</sup> for the 1<sup>st</sup> trimester time period (when only including prospective studies that used IgG avidity to determine timing of maternal infection and defined symptoms as severe sensorineural hearing loss and/or neurodevelopmental delay). We assume that the symptomatic risk for the pre/periconception period is the same as it is for the 1<sup>st</sup> trimester period.<br>cCMV: congenital cytomegalovirus; VT: vertical transmission; CI: Confidence Interval |  |  |

In the secondary comparisons, our calibrated input parameters were similar to the mean estimates from the literature. For *VT:primary risk*, our calibrated values ranged from 5.4 - 66.2%, depending on the timing of maternal infection, very close to the mean estimates of 5.5 - 66.2% from Chatzakis *et al*.^33^ For *maternal primary infection rate*, our calibrated value was 1.93/100PY, compared to the 1.95/100PY obtained from our analysis of the CDC NHANES data.^57,58^ For *symptomatic:primary*, our calibrated values ranged from 2.0-47.3%, depending on the timing of maternal infection, higher than the mean estimates of 0.6-21.8% derived from the studies reported in Chatzakis *et al.*^33^

The relative risk of vertical transmission from non-primary maternal infection compared to the risk from primary maternal infection (*RR-VT:non-primary*) was calibrated to be 0.495. Similarly, the relative risk of symptomatic cCMV for infant infections that follow non-primary maternal infection compared to those following primary maternal infection (*RR-symptomatic:non-primary*) was calibrated to be 0.584. Thus, for both vertical transmission and symptomatic risk, calibrated values for the risks following non-primary maternal infections were substantially lower than the risks following primary maternal infection.

## DISCUSSION

We developed a patient-level simulation model of CMV disease progression, testing, and treatment among mother-infant pairs during pregnancy. We tested and calibrated our modeling framework using published estimates from various cohort studies. The LINCS model presents several novel contributions to the CMV simulation modeling literature. First, the simulation of mother-infant dyads enables us to easily model interactions between maternal and fetal health states and allows for the incorporation of epidemiologic data for both maternal and fetal CMV outcomes during pregnancy. This not only results in more detailed and realistic simulation dynamics but also will facilitate analyses that identify cCMV testing and treatment strategies that are optimal for both the pregnant person and child. The LINCS model structure additionally provides flexibility in its ability to specify testing and treatment algorithms, the characteristics of diagnostic assays, and the effectiveness of therapies, allowing for comparison of the clinical impact of alternative testing and treatment strategies. This flexibility will allow the model to evaluate a range of potential clinical practices and guidelines, including in the context of comparative or cost-effectiveness analyses, to help guide clinicians and policymakers.

Data on many aspects of the biology of cCMV are limited, so we calibrated some particularly uncertain and influential parameters in our model. In our calibration analyses, the best-fitting parameter sets produced model output that matched the targets from the literature well (Table 3). Additionally, for the calibration to prevalence of cCMV at birth, we generated likelihood values representing the fit between the parameter sets and targets; in planned policy analyses using the LINCS model, these likelihood values can be used to conduct probabilistic sensitivity analyses that more fully capture the uncertainty in the available data.

Through the calibration, we found that the calibrated risks of VT after maternal primary infection match previously reported values, but that the proportion of cCMV infections that are symptomatic when they occur after maternal primary infection needed to be substantially higher than values previously reported in the literature for model output to match the calibration targets. We also found that the risks of vertical transmission and the risk of symptomatic cCMV following maternal non-primary infection were lower than those following maternal primary infection (relative risks of 0.495 and 0.584, respectively), but that these relative risks are higher than values often quoted in the literature.^29,59–61^ This may suggest that these underlying biologic risks are higher than the risks that are able to be observed in clinical trials or cohort studies, or that definitions of phenotypes used in clinical studies differ from those used in our model. Future analyses should conduct appropriate sensitivity analyses to assess the robustness of model output to changes in these parameters or potentially perform additional calibration if new targets become available.

### Limitations

This analysis has several limitations. The lack of comprehensive data for many of the model inputs required deriving data from several different studies, often in different geographic settings. Extrapolation of data between geographic settings was felt to be a reasonable assumption for biological parameters such as length of time of infection and test sensitivities and specificities, which are likely similar across regions and populations. In contrast, the calibration target for the prevalence of neonatal cCMV was drawn from a single US state (Minnesota) and the target for the proportion of symptomatic cases following cCMV infection was drawn from a US state and Canadian province (Minnesota and Ontario); these values might differ more substantially across populations.^46,62^ Future analyses may require further calibration with regional data to obtain appropriate and accurate results. Additionally, we do not explicitly model forms of transmission other than vertical. Transmission of CMV to pregnant people is modeled through a calibrated incidence value, not through dynamic transmission modeling. The current LINCS model is thus well-suited to address critical clinical policy questions about testing and treatment for mother-infant pairs, rather than the population-level impact of these strategies on all people who may transmit or acquire CMV or the impact of prevention strategies such as vaccination.^21,63^

## Conclusion

We report the development, testing, and calibration of the LINCS model of CMV infection during pregnancy. We have demonstrated that the model provides a reasonable simulation of risks of CMV vertical transmission, compared to estimates in the literature. We anticipate future expansions of the model to include details of prevention and treatment, as well as long-term infant outcomes, diagnosis, and care. LINCS will provide a framework that can be used in future work evaluating the clinical and economic impacts of prenatal CMV screening strategies, vertical transmission prevention methods, and novel treatments in development.

## Data Availability

This study used publicly available CMV seroprevalence data from the National Health and Nutrition Examination Survey (NHANES). All other data used in this paper is provided in the manuscript and supplemental material. The model code used to generate the data and results in the present study is available upon reasonable request to the authors.

https://wwwn.cdc.gov/nchs/nhanes/continuousnhanes/default.aspx

## SUPPLEMENTARY MATERIALS

### Priors for Bayesian Calibration

#### CMV Seroprevalence and Incidence

We estimated prior distributions for the CMV seroprevalence and primary incidence using data from three waves of the National Health and Nutrition Examination Survey (NHANES) collected from 1999-2004.^1^ The NHANES data for those years includes the results of CMV IgG antibody testing for all individuals 6 years of age and older. We estimated the age-specific CMV primary incidence rate by treating this cross-sectional seroprevalence data as current status data and estimating a hazard regression model with interval censoring for all female individuals.^2^ For seropositive individuals, the time of primary CMV infection is left censored, so that it is only known that the time of the initial CMV infection occurred between birth and the age the individual was screened for NHANES. For seronegative individuals, the time of primary CMV infection is right censored at the age of NHANES screening. Based on this interval censoring design, we fit a hazard regression using the *flexsurvspline* function in the *flexsurv* package for R – we included cubic splines for age with 3 knots in the regression, and boundary knots at birth and age 49 years.^3^ The internal knots were placed at equally spaced quantiles (i.e., 25%, 50%, 75%) of the midpoints for the interval between birth and the age of NHANES screening for all seropositive intervals (this approach is the default knot placement for the *flexsurv* package).

After fitting the hazard regression model, we extracted the estimated hazard rate at age 26 to parameterize the maternal primary CMV incidence rate in the simulation model. We also calculated the cumulative hazard between birth and age 26 to estimate maternal CMV seroprevalence at model start. To estimate the non-primary CMV incidence rate, we used data from Paris et al.^4^, which reported the results of a prospective cohort study of adolescent girls – out of 146 and 204 seronegative and seropositive participants, there were 22 primary and 12 non-primary (defined as >4-fold IgG increase) infections over 36 months. In order to be able to draw from a distribution for the relative risk of non-primary to primary infection, we fit a Poisson regression with a log link function (implemented with the *glm* function in R) with an offset term for the number of individuals seropositive or seronegative at baseline to calculate the relative risk of non-primary to primary infection:

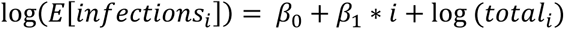

Here, *i* is 0 for the baseline seronegative group and 1 for the baseline seropositive group, *infection*_*i*_ is the number of observed infections after baseline in group *i*, *total*_*i*_ is the number of individuals in group *i*, *β*_0_ is an intercept term capturing the infection risk for the baseline seronegative group (on the log-risk scale), and *β*_1_is the log-relative risk for the baseline seropositive group (compared to the seronegative group). We multiplied the primary CMV incidence rate by this relative risk (exp (*β*_1_)) to estimate the non-primary incidence rate at age 26.

To generate draws from the prior distribution for CMV seroprevalence and incidence rates for use in the Bayesian calibration process, we used a bootstrap approach for the estimates from the hazard regression to account for the complex survey design in NHANES. We generated 10,000 replicate datasets using the *svrep* R package,^5^ which uses the sampling method described by Beaumont and Émond.^6^ Within each replicate, we estimated the CMV seroprevalence and primary incidence rate at age 26 using the hazard regression approach described above. We also drew a sample for the relative risk of non-primary to primary incidence within each replicate by assuming a normal distribution for the log relative rate, with the mean and standard deviation set based on the estimated log relative rate and associated standard error from the coefficient for the Poisson regression described above (i.e., *β*_1_). We multiplied this draw for the relative risk by the estimated primary incidence rate within the replicate to generate a draw for the non-primary incidence rate at age 26.

#### Vertical Transmission Risk and Symptomatic cCMV Infection Risk

We estimated the prior distributions for the vertical transmission risk and symptomatic cCMV infection risk following maternal primary infection (stratified by the timing of the infection) using the studies reported by Chatzakis *et al.*^7^ We used the studies reported in Figure 2 and Supplemental Figure 7 from Chatzakis *et al.*^7^ for vertical transmission and symptomatic infection risk, respectively. For each of the studies, we extracted the sample size (i.e., mother-infant pairs, cCMV infections) and number of observed events (i.e., vertical transmission, symptomatic cCMV infection) across the different timings for primary maternal infection, and then fit a binomial regression with a logit link and study-level random effects:

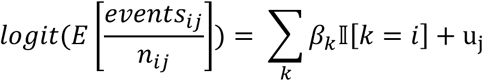

Here, *events*_*ij*_ is the number of observed events in infection timing group *i* (e.g., maternal infection in first trimester) in study *j*, *n*_*ij*_ is the sample size for the relevant timing group and study, *β*_*k*_ is the log-odds of the event for infection timing group *k*, and *u*_*j*_ is the study-level random effect (assumed to have a normal distribution with mean 0). This regression-based approach to the meta-analysis of risks is analogous to the Poisson regression approach recommended by Spittal et al.^8^ and Böhning et al.^9^ We used a Bayesian approach to fit these regressions, implemented with the *rstanarm* R package;^10^ we used the default weakly informative priors and 100,000 iterations across 4 chains to estimate the model. After fitting the model, we used 10,000 samples as the prior distribution for the vertical transmission and symptomatic cCMV risk across the different timings of maternal primary infection. To estimate the vertical transmission and symptomatic cCMV risk for non-primary maternal infection, we assumed a uniform prior distribution between 0-1 for the relative risk of these events comparing non-primary to primary maternal infection – we took two sets of 10,000 draws from the uniform prior (one each for the vertical transmission and symptomatic risks) and multiplied it by the samples for the risks after a primary maternal infection to generate the draws for vertical transmission and symptomatic cCMV risk following non-primary maternal infection.

### Unit testing and functional testing of model dynamics

Code testing is typically done by checking that model code produces expected outputs when provided with a specified set of inputs. Given the complex and often nonlinear nature of simulation models, however, it is often not straightforward or possible to calculate the expected outputs explicitly. To address this, we adapt a testing paradigm from software engineering that we refer to as *unit testing* and *functional testing*, which are code validation methods that ensure that the code works as specified. Unit testing involves testing individual functions or portions of code and can help identify structural and logical errors in the code. For deterministic (e.g., structural) unit testing, conditional responses were added in key functions that automatically returned errors if unreasonable or nonsensical events occurred. This prevented potential errors from propagating through the rest of the model before being caught, which would have increased the complication of diagnosing and fixing the issues. For example, in the section of the code that draws from a probability for vertical transmission of CMV infection to the fetus, we created a conditional response that forced the model to return an error warning if vertical transmission occurred without previous maternal CMV infection. Unit testing was also performed to ensure that events were occurring at the rate at which they were specified, such as making sure that the sensitivities and specificities of the simulated prenatal tests matched their input values.

Functional tests, in contrast to unit tests, are passed through multiple functions and model parts to evaluate performance across linked or nested functions, testing to see whether the integrated system behaves as expected in various scenarios. An example of a functional test that was performed entailed confirming that the number of each prenatal test performed appropriately correlated with the user-defined proportion of active maternal infections that are symptomatic and the proportion of symptomatically infected individuals who seek healthcare. Functional testing allowed us to verify that increasing the proportion of symptomatic CMV infections, increasing the proportion of pregnant people with symptoms who seek healthcare, and increasing the proportion of those seeking healthcare who get tested for CMV all directly correlate to a greater number of maternal CMV cases diagnosed, which is logically to be expected given our model structure. A schematic of this test is outlined in Figure S1.

**Figure S1.**
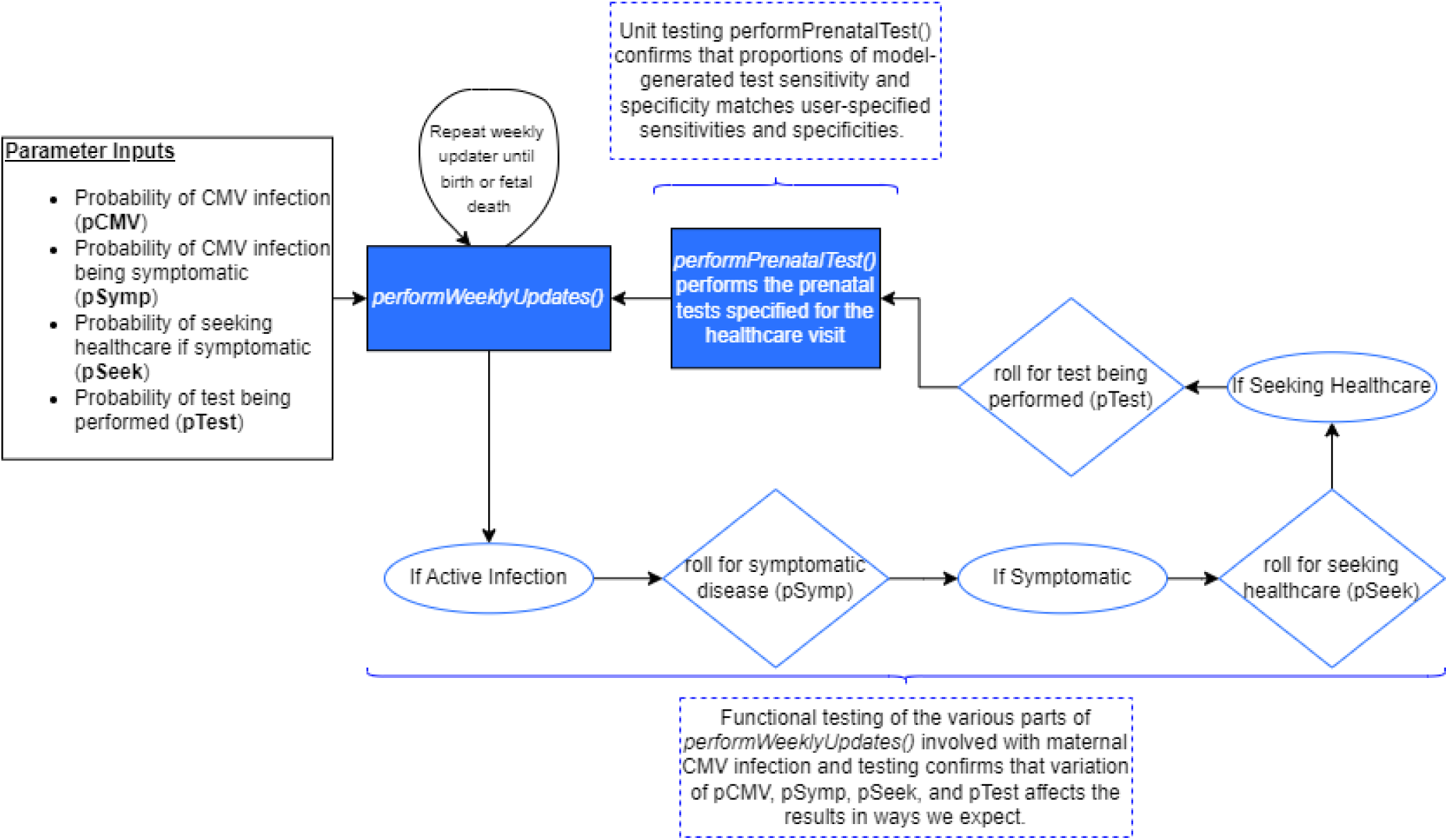
Unit and functional testing. In this simplified and noncomprehensive model schematic, we compare examples of unit and functional testing that were performed.

**Table S1.** Results of functional testing: Sample inputs and intermediate outcomes.

| Parameter | Target value | Observed value | Relative difference | Approach to tracking |
| --- | --- | --- | --- | --- |
| IgM sensitivity and specificity | 90%, 96% | 89.8%, 95.99% | <1%, <1% | The model code tracks the total number of true positives, false positives, true negatives, and false negatives for each test. These values were then used to calculate the observed sensitivity and specificity for each prenatal test. |
| IgG sensitivity and specificity | 97%, 96% | 97.06%, 95.99% | <1%, <1% |  |
| IgG avidity sensitivity and specificity | 94.3%, 100% | 94.32%, 100% | <1%, <1% |  |
| Serum CMV DNA PCR sensitivity and specificity | 94%, 99% | 93.99%, 98.999% | <1%, <1% |  |
| Amniotic fluid CMV DNA PCR sensitivity and specificity | 86%, 100% | 85.6%, 100% | <1%, <1% |  |
| Number of tests performed | N/A |  |  | It was verified that increasing the proportion of infected mothers who are symptomatic, increasing the number of symptomatic mothers who seek healthcare, and increasing the probability of those who seek healthcare getting tested all resulted in more prenatal tests being performed. This allowed us to ensure that all of these interrelated parts of the model were functioning together properly. |
| Other structural checks | N/A |  |  | Model run is stopped and error is displayed if:<br>(1) Test results return any value that is not either true (1) or false (0),<br>(2) A test that is not fully specified by the user is called upon. |

